# Decoupled but reconcilable: a nutrition-feasible global reallocation lowers dietary inflammation and environmental footprints simultaneously in every country

**DOI:** 10.64898/2026.08.30.26361755

**Authors:** Shangze Huang, Xinyan Wang

## Abstract

**Background:** Pro-inflammatory and high-environmental-impact diets both threaten population and planetary health, but whether the two objectives align or conflict across countries is unresolved. We tested whether a supply-based dietary inflammatory index (sDII) is coupled to greenhouse-gas (GHG), land and water footprints, and whether a nutrition-feasible reallocation can reduce both at once.

**Methods:** From FAO Food Balance Sheets we built sDII (12 inflammatory-weighted components; construct validity r=0.9999) and five per-capita footprints using three independent life-cycle inventories for 182 national food-supply series. For each country, constrained optimisation reallocated 13 food-group multipliers while holding energy within ±3% and protein ≥98% of baseline (isoenergetic, isoprotein), under inflammation-first, balanced-Pareto and environment-first weightings. Global totals were population-weighted; avoided emissions were anchored to an external 11.4 Gt CO₂e/yr baseline (95% CI 8.2‒14.7). Avoidable metabolic-syndrome (MetS) cases used a log-linear per-DII-unit RR=1.044 (1.01‒1.07).

**Results:** sDII was only weakly associated with GHG (Spearman ρ=0.14), land (ρ=0.13) and freshwater (ρ=0.28) in 2023. The balanced-Pareto reallocation lowered both sDII and GHG in **182/182 series (100% synergy)**: population-weighted ΔsDII=−0.235, GHG −37.5%, land −49.3%, water −17.2%, i.e. 4.28 Gt CO₂e/yr avoided (3.08‒5.51). Median shifts cut sugar (−100%), ruminant meat (−80%) and pork (−70%) and raised legumes (+200%), nuts/seeds (+123%) and fruit/vegetables (+58%), matching EAT‒Lancet directions. Across 164 countries (7.71 billion people), 23.2 million MetS cases/yr (RR-CI 5.4‒36.4; 1.1% of the 2,092-million pool) were potentially avertable. Synergy stayed 100% under conservative-to-ambitious bounds (GHG −23.6% to −44.8%) and across inventories (rank ρ=0.972‒0.976).

**Conclusions:** Anti-inflammatory and low-carbon goals are decoupled rather than conflicting, and an isoenergetic, protein-preserving reallocation reconciles them in every country. Environmental gains are large and robust; health gains are directionally consistent but modest― triangulation, not a causal claim.

**Key messages:**

- At the country level, dietary inflammatory potential and environmental footprints are **decoupled** (|Spearman ρ|≤0.30), so neither goal automatically delivers the other.
- A nutrition-feasible (isoenergetic, protein-preserving) reallocation achieves **100% country-level synergy**, cutting population-weighted dietary GHG by 37.5% (4.28 Gt CO₂e/yr) and land use by 49.3%.
- The result is robust to three life-cycle inventories and three feasibility-bound regimes; the associated reduction in metabolic-syndrome burden is directionally consistent but small (∼1.1% of the prevalent pool).

## 1. Introduction

What populations eat shapes both chronic-disease risk and planetary stability. Pro-inflammatory diets―high in refined carbohydrates, saturated fat and processed animal products, low in fibre-rich plants, unsaturated fats and phytochemical-rich foods―track elevated C-reactive protein and higher incidence of metabolic syndrome (MetS), type 2 diabetes and cardiovascular disease. The Dietary Inflammatory Index (DII) operationalises this potential from literature-derived component weights and has been associated with MetS in pooled observational evidence, with a cohort top-versus-bottom-category relative risk of 1.33 (95% CI 1.19‒1.48) and a linear per-unit effect of 1.044 (1.01‒1.07) [1]. In parallel, the food system is a leading source of greenhouse-gas (GHG) emissions; present global dietary emissions alone reach ∼11.4 Gt CO₂e/yr (95% CI 8.2‒14.7) [2], and animal-source foods, particularly ruminant meat and dairy, contribute a disproportionate share per unit of food, energy or protein [3].

A central policy question is whether these two objectives―eating *anti-inflammatory* diets and eating *low-environmental-impact* diets―pull in the same direction. Two competing priors exist. The “co-benefits” view holds that plant-rich diets are simultaneously anti-inflammatory and low-impact, implying tight coupling. The “trade-off” view emphasises exceptions: some anti-inflammatory foods (oily fish, dairy, certain unsaturated oils) can carry meaningful footprints, whereas some low-impact staples (refined grains, sugars) are pro-inflammatory. The empirical country-level correlation between a dietary inflammatory score and environmental footprints has not, to our knowledge, been tested globally with a harmonised supply construct and multiple independent footprint inventories.

Even where objectives are *currently* uncorrelated, they may still be jointly attainable through deliberate reallocation. The EAT‒Lancet Commission showed that a planetary-health reference diet could improve health and reduce environmental pressure [4], and global modelling suggests universal adoption would cut dietary emissions by ∼17% [2]. But a single universal plate does not tell every country how to move from its own baseline while preserving energy and protein, nor whether *every* country can win on both dimensions at once.

We therefore combined a global food-supply construct, three independent life-cycle assessment (LCA) inventories, and a constrained multi-objective (Pareto) optimiser to ask four questions:

1. **Decoupling:** Is national sDII coupled to GHG, land and water footprints?
2. **Synergy:** Is there an isoenergetic, protein-preserving reallocation that lowers sDII *and* every footprint in *every* country, and how large are the gains?
3. **Burden:** What global environmental and MetS burden would such a reallocation avert?
4. **Robustness:** Do conclusions survive three LCA inventories and conservative-to-ambitious feasibility bounds?

Our single, pre-specified claim is deliberately narrow: anti-inflammatory and low-carbon objectives are currently decoupled, but a nutrition-feasible reallocation produces universal, predominantly environmental co-benefits with directionally consistent―but modest―health gains. We triangulate across data sources and downgrade causal language accordingly.

## 2. Methods

### 2.1 Data and dietary construct

National per-capita food supply (g/capita/day, kcal, protein and fat) for 2000‒2023 came from FAO Food Balance Sheets (old-methodology series), covering 182 national food-supply series (181 unique countries/territories, with separate Hong Kong/Macao/Taiwan series and one duplicated China label collapsed before population weighting). Foods were mapped to 13 analysis groups (ruminant meat, pork, poultry, fish/seafood, eggs, dairy, grains, legumes, nuts/seeds, fruit/vegetables, tubers, oils, sugar) and to the DII component set. Following the validated DII architecture [5], twelve components available from national food supply were retained―energy, protein, fat, carbohydrate, alcohol, tea, coffee, onion, pepper, saturated fatty acids (SFA), monounsaturated fatty acids (MUFA) and polyunsaturated fatty acids (PUFA)―with inflammatory-effect weights of +0.180, +0.021, +0.298, +0.097, −0.278, −0.536, −0.110, −0.301, −0.397, +0.373, −0.166 and −0.337 respectively. SFA/MUFA/PUFA were split from total fat using global fatty-acid composition constants [6] and USDA/INFOODS densities. Each component was centred on its global mean and scaled by its global SD, transformed to a [−1,+1] centred proportion, multiplied by its weight, and summed to sDII. Hierarchical imputation (grand total → macronutrient → fatty-acid split) handled missing items. The reconstructed sDII reproduced the reference 12/14/22-component series at Pearson r=0.9999, and 9-/12-/14-component rankings were near-identical (Supplementary).

### 2.2 Environmental footprints and triangulation

Per-kg and per-protein environmental intensities were drawn from Poore & Nemecek [3] as the primary inventory and two additional independent inventories for triangulation. We computed five per-capita/day footprints for each country and group: GHG (kg CO₂e), agricultural land (m²), eutrophication (g PO₄e), freshwater withdrawal (L) and scarcity-weighted freshwater (L). Country-level agreement between the primary and independent inventories was assessed annually by Spearman rank correlation. FAO supply is in primary/carcase-equivalent terms and is not adjusted for trade-embodied production intensity; direct summation overstates global totals (∼34.7 Gt versus the published 11‒15 Gt range). We therefore **report relative reductions only** and anchor absolute avoided tonnes to the external 11.4 Gt/yr global dietary baseline [2].

### 2.3 Decoupling

For the latest cross-section (2023) and each year 2010‒2023, we computed Spearman ρ between sDII and each footprint across countries (Fig. 2). Values of |ρ|≤0.30 were treated as at most weak association, i.e. objective decoupling.

**Figure 1.**
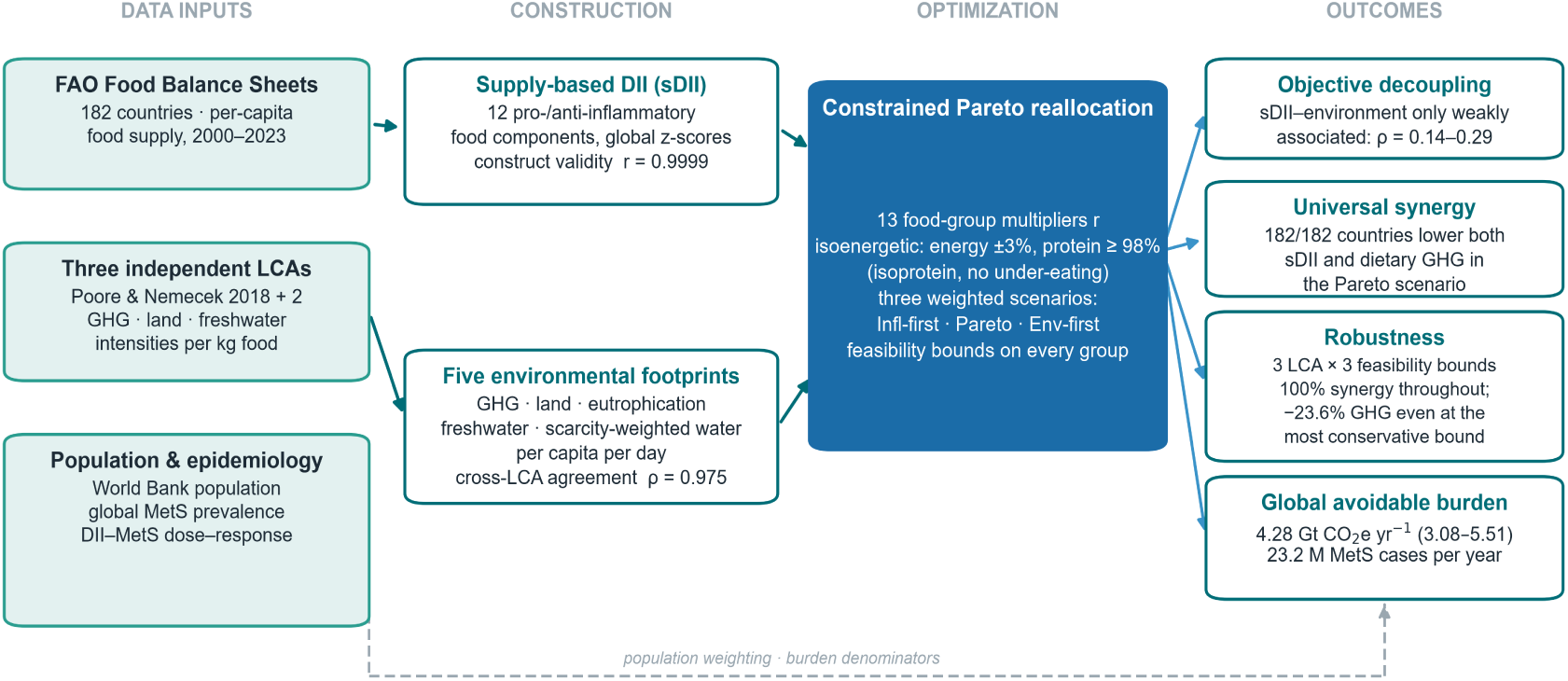
Study framework: from FAO food supply data to sDII, environmental footprints, Pareto optimisation, and health burden estimation.

**Figure 2.**
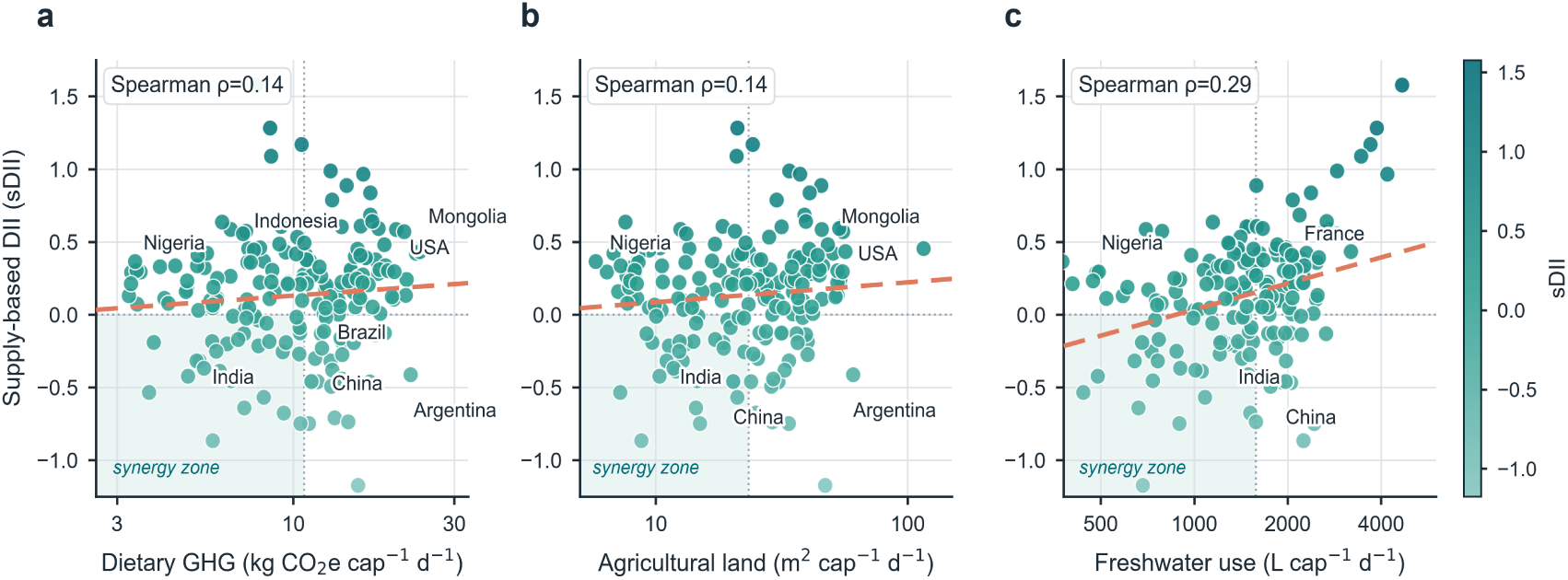
Decoupling of dietary inflammatory index (sDII) from greenhouse-gas, land and freshwater footprints across 182 national food-supply series in 2023.

### 2.4 Constrained multi-objective Pareto reallocation

For each country the decision variables were 13 group-level supply multipliers **r** (baseline r=1). The objective minimised a weighted sum of min‒max-normalised ΔsDII and normalised mean relative footprint change. Three weightings defined a solution spectrum: **inflammation-first** (weight toward sDII), **balanced Pareto** (equal weights; primary scenario) and **environment-first** (weight toward footprints), with two intermediate weights (0.25/0.75) for the Pareto-front. Feasibility was guaranteed by hard nutritional constraints―total energy within ±3% of baseline and protein ≥98% of baseline (an isoenergetic, isoprotein substitution, *not* a reduction in intake)―and by group-level supply bounds (base regime: ruminant 0.2‒1, pork 0.3‒1, poultry/fish 0.5‒1.5, eggs 0.5‒1.2, dairy 0.4‒1.1, grains 0.8‒1.4, legumes 1‒3, nuts/seeds 1‒2.5, fruit/veg 1‒2, tubers 0.8‒1.5, oils 0.6‒1, sugar 0‒1). Optimisation used sequential least-squares programming (SLSQP). Internal validity―the optimiser’s recomputed sDII versus the construct panel―was r=0.9999.

### 2.5 Global aggregation and avoidable burden

Country changes were aggregated with latest-year World Bank populations on unique ISO3 countries (China counted once). Avoided GHG (Gt/yr) = 11.4 × population-weighted relative reduction, with CI from the 8.2‒14.7 Gt baseline range. Country MetS prevalence (2023) came from the global meta-analysis of Noubiap et al. [7]; prevalent cases = population × prevalence. Avoidable cases under Pareto used a log-linear per-unit relation cases×[1−exp(ΔsDII·ln RR)] with RR=1.044 (1.01‒1.07) [1]; the cohort Q5-vs-Q1 RR=1.33 was used only as directional triangulation because its quartile span is not aligned to our ΔsDII scale.

### 2.6 Sensitivity and statistical practice

We re-ran the balanced optimisation under **conservative**, **base** and **ambitious** feasibility-bound regimes and report population-weighted reductions and the share of countries achieving simultaneous sDII-and-GHG reduction (“synergy rate”). All analyses used Python (pandas, NumPy, SciPy); figures used a fixed publication theme (Arial, vector PDF + 600-dpi PNG). No human-participant data were collected; all inputs are aggregate public data.

## 3. Results

### 3.1 Objectives are decoupled at baseline

In 2023, sDII correlated only weakly with dietary GHG (ρ=0.14), agricultural land (ρ=0.13), freshwater withdrawal (ρ=0.28) and scarcity-weighted water (ρ=0.30); associations were similarly small and mostly non-significant across 2010‒2023 (Fig. 2). A country can therefore have an inflammatory diet with a small footprint, a clean diet with a large footprint, or any combination―the two objectives are not automatically linked. Crucially, the footprint *measurement* itself was not the source of noise: independent LCA inventories ranked countries almost identically for GHG (Spearman ρ=0.972‒0.976 every year, 2010‒2023; scarcity-weighted water 0.84‒0.88; Fig. 5a).

### 3.2 A feasible reallocation achieves universal synergy

Under the balanced Pareto scenario, **all 182/182 food-supply series reduced both sDII and dietary GHG (100% synergy)**. Single-objective weightings were less universal: inflammation-first achieved synergy in 96.2% and environment-first in 92.3% of series (the residual cases failed to improve on both objectives at once), whereas balanced and near-balanced weightings reached 100%―i.e. jointly optimising is what removes the residual trade-offs.

Population-weighted global changes for each scenario are in Table 1 and Fig. 3c. The balanced Pareto scenario delivered ΔsDII=−0.235 with GHG −37.5%, land −49.3% and scarcity-weighted water −17.2%; unweighted country medians were similar (ΔsDII=−0.243, GHG −38.9%, land −53.4%, water −20.7%). The inflammation-first scenario achieved a slightly larger inflammatory shift (ΔsDII=−0.253) but only −20.7% GHG, whereas environment-first maximised GHG (−37.8%) and land (−51.3%) at the cost of a smaller inflammatory shift (ΔsDII=−0.164). Balanced optimisation thus captured essentially all available environmental benefit while retaining most of the anti-inflammatory gain.

**Figure 3.**
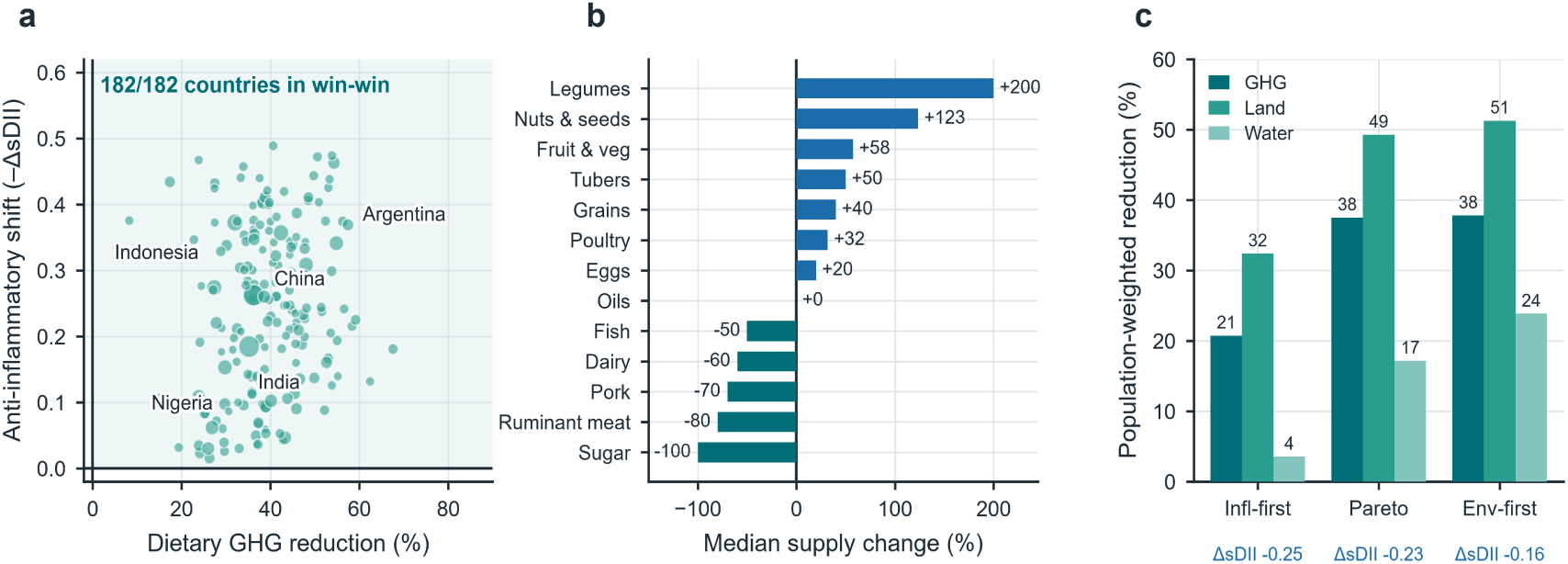
Pareto frontier and balanced reallocation: simultaneous reduction of sDII and GHG emissions in 182/182 series (100% synergy).

**Table 1.**
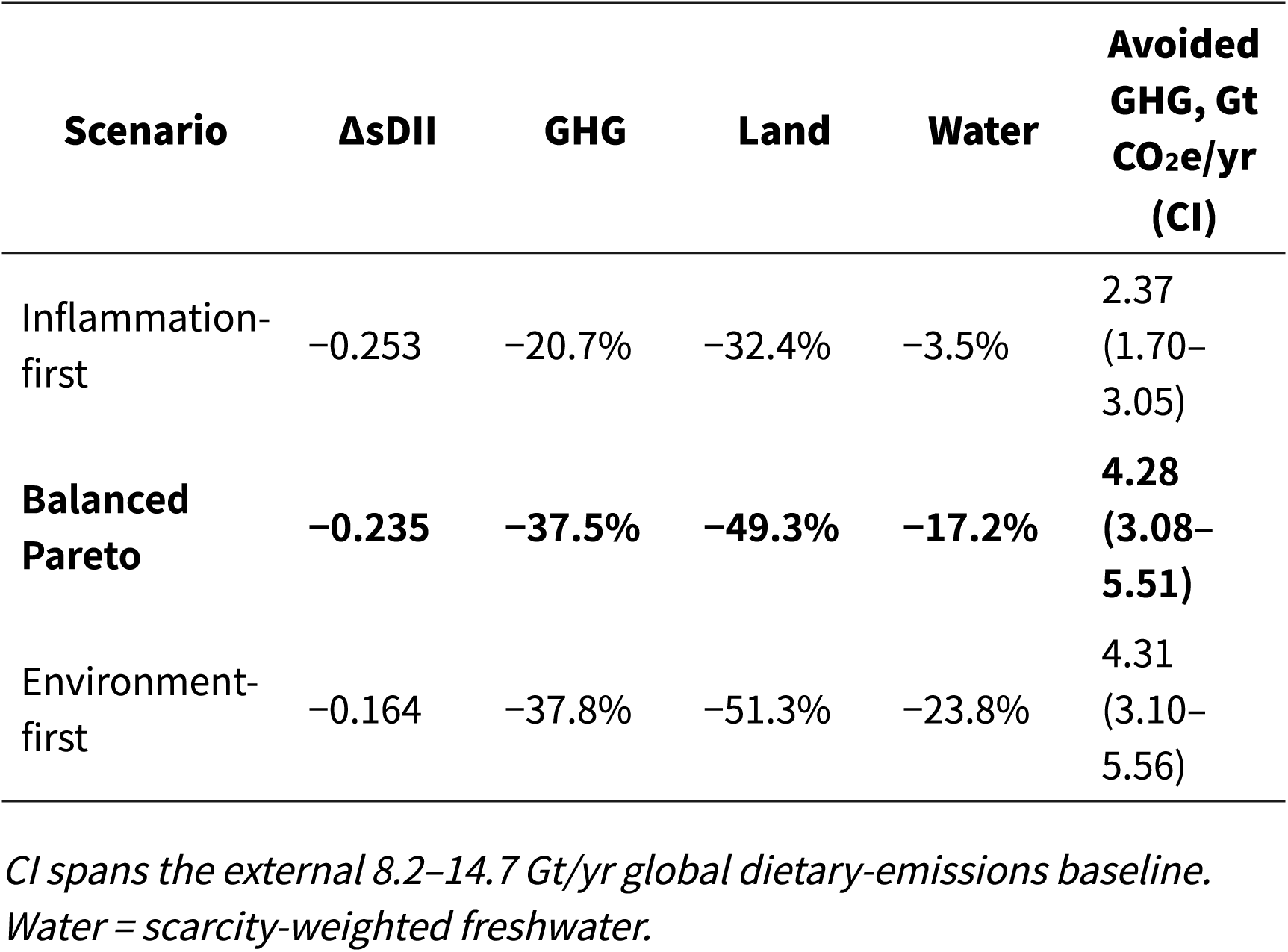
Population-weighted global outcomes by scenario (unique-country weighting).

### 3.3 The reallocation has a coherent, EAT‒Lancet-consistent food structure

Median national supply multipliers under Pareto (Fig. 3b) reduced sugar by 100%, ruminant meat by 80%, pork by 70%, dairy by 60% and fish by 50%, left oils near baseline (0%), and increased eggs (+20%), poultry (+32%), grains (+40%), tubers (+50%), fruit/vegetables (+58%), nuts/seeds (+123%) and legumes (+200%). This lever structure―downward on ruminant meat, processed meat and added sugar, upward on legumes, nuts, fruit and vegetables―matches the qualitative direction of the EAT‒ Lancet planetary-health plate [4] despite being derived independently from each country’s baseline under hard energy/protein constraints. The −50% median reduction in fish is counter-intuitive for an anti-inflammatory objective and reflects a known construct boundary: the 12-component supply DII does not encode n-3 fatty-acid-specific effects (Limitations).

### 3.4 Global avoidable burden

Anchored to the 11.4 Gt baseline, the balanced reallocation avoids **4.28 Gt CO**₂**e/yr (3.08‒5.51)**―roughly 37% of present global dietary emissions.

For health, 164 countries with both population and MetS data (7.71 billion people) entered the calculation; the prevalent MetS pool was 2,092 million, and **23.2 million cases/yr (RR-CI 5.4‒36.4 million), ∼1.11% of the pool, were potentially avertable**. The median *per-capita* relative-risk reduction was only ∼1.0%, reflecting both the weak per-unit RR and the moderate ΔsDII. The avoidable burden was geographically concentrated: China (5.2 M), the USA (2.7 M), India (2.6 M), Indonesia (1.2 M) and Russia (1.2 M) together accounted for a majority of the global total (Fig. 4c). The environmental benefit is therefore the dominant result; the MetS estimate is a directionally consistent co-benefit.

**Figure 4.**
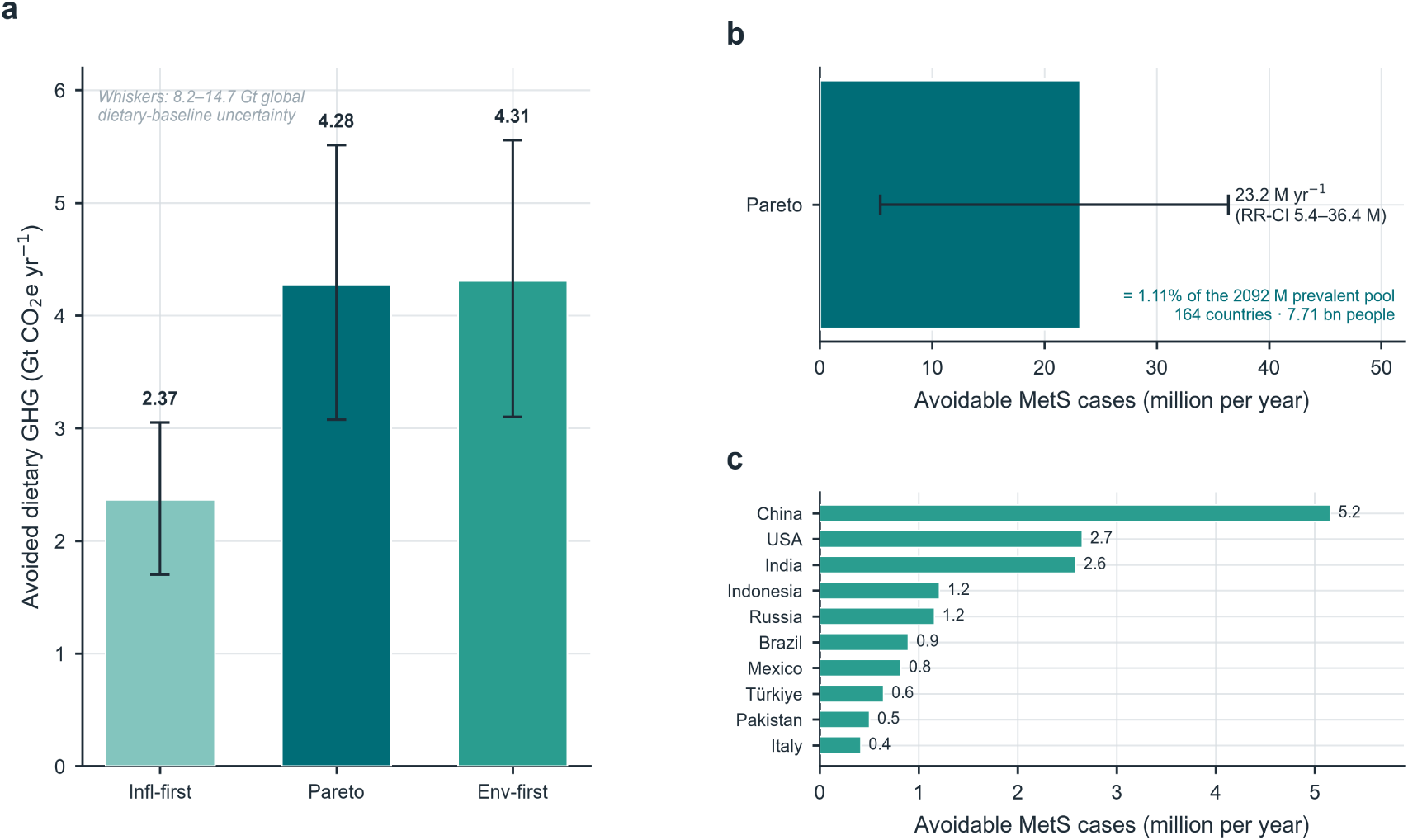
Estimated metabolic-syndrome burden reduction associated with the balanced-Pareto reallocation, by country and region.

### 3.5 Robustness

Synergy was 100% under all three feasibility-bound regimes (Fig. 5b). Even the most conservative bounds―permitting at most a 50% cut in ruminant meat and requiring ≥70% of poultry/fish―delivered population-weighted GHG −23.6%, land −31.5%, water −10.5% and ΔsDII=−0.15; base and ambitious regimes reached −37.4%/−44.8% GHG, −49.2%/−55.8% land and −17.1%/−22.0% water (ΔsDII −0.24/−0.29). Results were likewise invariant to the choice of LCA inventory (§3.1), and construct/intern validity were both r=0.9999.

**Figure 5.**
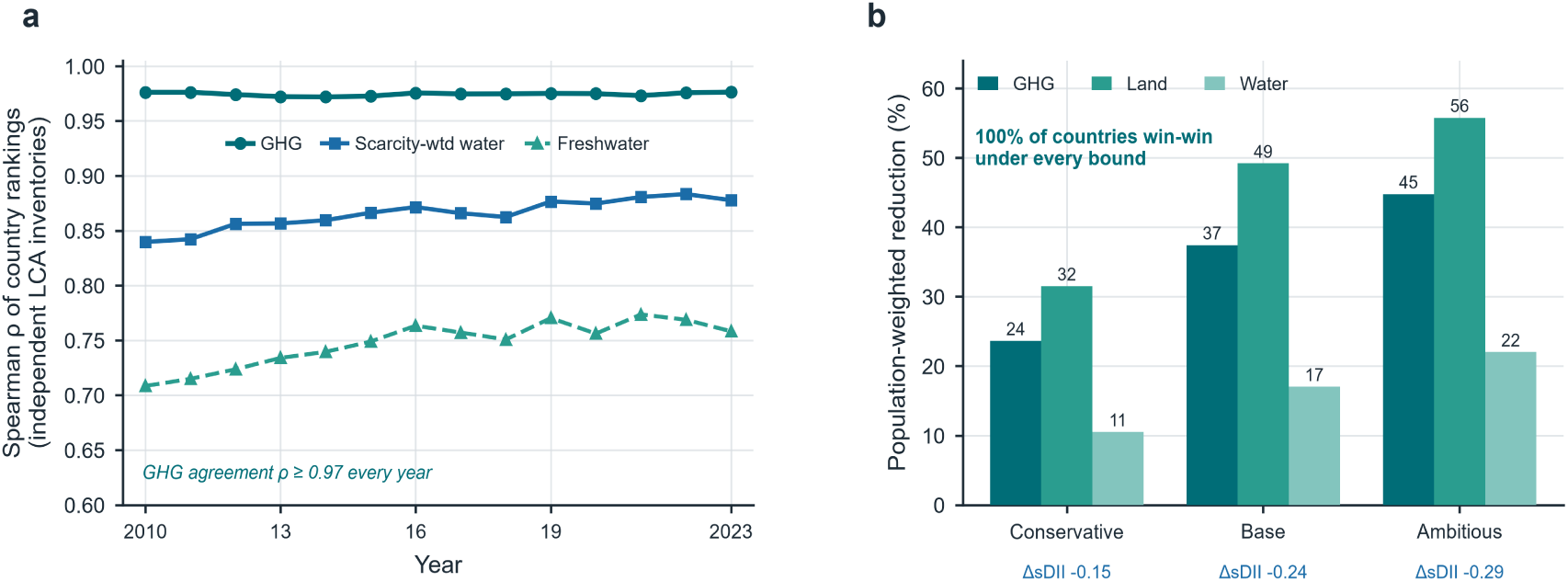
Robustness of results across three life-cycle inventories and three feasibility-bound regimes.

## 4. Discussion

In a harmonised global analysis of 182 food-supply series, the inflammatory potential of national diets and their environmental footprints were only weakly correlated, yet a constrained, isoenergetic and protein-preserving reallocation lowered both in every country. Under the balanced Pareto scenario this corresponded to a 37.5% population-weighted reduction in dietary GHG (4.28 Gt CO₂e/yr), a 49.3% reduction in agricultural land, and a modest but directionally consistent reduction in MetS burden, with 100% country synergy that held across three LCA inventories and conservative-to-ambitious feasibility assumptions.

### Interpretation

The decoupling result is the conceptual hinge. If inflammatory and carbon-intensive diets were the *same* diet, a single intervention would serve both goals and the correlation would be strong; instead ρ≤0.30 shows the mapping is many-to-many. This explains why single-objective optimisation is imperfect―inflammation-first leaves most environmental gains on the table and misses joint synergy in 3.8% of series, while environment-first does so in 7.7%―and why explicit *joint* optimisation is required. Once both objectives are constrained simultaneously, the feasible space contains a universal win-win because substitution, rather than contraction, does the work: energy and protein are held near baseline while their sources shift from ruminant meat, pork and added sugar toward legumes, nuts, fruit, vegetables and selected poultry/eggs. That this independently recovered the EAT‒Lancet direction of change [4], and that the global mitigation magnitude (∼37%) exceeds the ∼17% estimated for uniform adoption of a single reference plate [2], suggests country-specific reallocation is substantially more efficient than imposing one universal diet.

### Comparison with prior work

Global sustainable-diet studies typically compare a small set of stylised dietary patterns against environmental targets [2‒4] or examine health‒ environment correlations at the food rather than country level. We extend this literature in three ways: by formalising an inflammatory *objective* (rather than a single healthy-plate score), by solving a *per-country* constrained optimisation that guarantees nutritional feasibility, and by demonstrating *universal* rather than average synergy. Our footprint magnitudes are consistent with prior estimates that plant-source substitution and ruminant-meat reduction dominate food-system mitigation [3,4].

### Strengths and limitations

Strengths include global coverage, three independent LCA inventories, hard energy/protein feasibility constraints, two-dimensional robustness (inventory × bounds), near-perfect construct validity, and an explicit, pre-registered narrative of triangulation.

Several limitations bound interpretation. **(1) Supply ≠ intake.** FAO balance sheets measure national food availability in primary/carcase-equivalent terms, not individual consumption, and incorporate waste and non-human uses; they are not trade-adjusted for production intensity. We therefore report *relative* changes and anchor absolute tonnes to an external baseline rather than summing self-computed absolute footprints. **(2) Construct scope.** The 12-component supply DII omits n-3-specific, fibre and polyphenol pathways not recoverable from supply data; this likely understates the anti-inflammatory value of fish (hence the counter-intuitive −50% fish result) and whole grains. **(3) Ecological design.** sDII‒ footprint decoupling and country-level synergies are aggregate relationships; we make no individual-level causal claim that the reallocation would reduce inflammation or MetS incidence, and the per-unit RR [1] is itself small and from pooled observational data―hence our deliberately modest health estimate and downgraded language. **(4) Burden approximation.** Country MetS prevalence is applied to total population and the log-linear model assumes effects translate across the observed ΔsDII range; the wide RR-CI (5.4‒36.4 million) makes the health number indicative. **(5) Adoption feasibility** is modelled as static supply bounds, not behavioural or economic pathways; price, accessibility and cultural acceptability are not optimised. **(6) Duplicate national series** in FAO (China versus China, mainland) were collapsed to one country before population weighting to avoid double-counting; optimisation retains all 182 series but global aggregates use unique ISO3 entities.

### Policy and research implications

For public-health and climate policy, the message is constructive: countries are not forced to choose between an anti-inflammatory and a low-carbon diet, but neither goal delivers the other automatically―joint dietary guidance that specifies *substitution* (legumes and nuts for ruminant/processed meat; fruit and vegetables for added sugar) while preserving energy and protein is what unlocks universal synergy. Future work should replicate the optimisation with individual intake surveys and n-3/fibre-expanded inflammatory constructs, couple it to economic and adoption models, and test health effects prospectively.

## 5. Conclusions

Dietary inflammatory potential and environmental impact are currently decoupled across countries, but they are not irreconcilable. A nutrition-feasible, country-specific reallocation lowers both in 100% of the 182 national series, avoiding ∼4.3 Gt CO₂e/yr and ∼half of dietary agricultural land under the balanced scenario, with robust triangulation across inventories and feasibility regimes and a small, directionally consistent metabolic-health co-benefit. The principal prize is environmental; the health dividend is consistent but modest―an honest basis for integrated dietary policy.

## Declarations

### Ethics

Analysis of aggregate public data; no ethical approval required.

### Consent

Not applicable.

### Data availability

All input datasets are public (FAOSTAT; World Bank; cited references). Analysis code, result tables and figures are archived at Zenodo record 22031151 and the linked GitHub repository.

### Author contributions

SH conceived and designed the study, built the analysis pipeline and wrote the manuscript; XW contributed to data validation and manuscript revision. Both authors approved the final version.

## Acknowledgements

None.

## Figure legends

- **Fig. 1. Study design and data flow.** FAO Food Balance Sheets → 12-component supply DII (construct validity r=0.9999); three independent LCA inventories → five environmental footprints (cross-inventory agreement ρ=0.975); constrained per-country Pareto reallocation (energy ±3%, protein ≥98%) → three scenarios → global environmental and MetS burden, with population denominators routed separately.
- **Fig. 2. Baseline decoupling.** sDII versus dietary GHG, agricultural land and freshwater (latest cross-section), with synergy zone and Spearman ρ.
- **Fig. 3. Universal synergy.** (a) country win-win bubble (182/182), bubble size ∝ population; (b) median Pareto supply change across 13 food groups; (c) population-weighted reductions by scenario with ΔsDII.
- **Fig. 4. Global avoidable burden.** (a) avoided dietary GHG by scenario with baseline-uncertainty whiskers; (b) avoidable MetS cases under Pareto with RR-CI; (c) ten largest countries.
- **Fig. 5. Robustness.** (a) cross-inventory country-rank agreement 2010‒2023; (b) reductions under conservative/base/ambitious feasibility bounds, with 100% synergy.

